# Aging-related changes in body composition across the lifespan – insights from over 66.000 individuals of a Western European population

**DOI:** 10.1101/2025.05.23.25328227

**Authors:** Matthias Jung, Marco Reisert, Hanna Rieder, Susanne Rospleszcz, Tobias Haueise, Tobias Pischon, Thoralf Niendorf, Hans-Ulrich Kauczor, Henry Völzke, Robin Bülow, Maximilian F. Russe, Christopher L. Schlett, Michael T. Lu, Fabian Bamberg, Vineet K. Raghu, Jakob Weiss

## Abstract

Body composition (adiposity and muscle depots) is strongly associated with cardiometabolic risk. However, using body composition measures for future disease risk prediction is difficult as they may reflect total body size or typical aging rather than poor health. We used data from the UK Biobank (UKB) and the German National Cohort (NAKO) to calculate age-, sex-, and height-specific z-scores for body composition measures (subcutaneous adipose tissue (SAT), visceral adipose tissue (VAT), skeletal muscle (SM), SM fat fraction (SMFF), and intramuscular adipose tissue (IMAT)) and describe changes across the lifespan. Multivariable Cox regression assessed the prognostic value of z-score categories (low: z<-1; middle: z=-1 to 1; high: z>1) for incident diabetes, major adverse cardiovascular events (MACE), and all-cause mortality beyond traditional cardiometabolic risk factors in the UKB. Among 66,608 individuals (mean age: 57.7±12.9 y; mean BMI: 26.2±4.5 kg/m^2^, 48.3% female), SAT, VAT, SMFF, and IMAT were positively, and SM negatively associated with age. In multivariable-adjusted Cox regression, z-score risk categories had hazard ratios of up to 2.69 for incident diabetes (high VAT category), 1.41 for incident MACE (high IMAT), and 1.49 for all-cause mortality (low SM) compared to middle categories. Body composition shows distinct age-related changes across the lifespan. Z-scores of age-, sex-, and height-adjusted body composition measures identify individuals at risk and predict cardiometabolic outcomes and mortality beyond traditional risk factors. Our open-source tool facilitates the clinical translation of age-specific body composition assessments and supports future research.

**One Sentence Summary:** Age-, sex-, and height-adjusted body composition z-scores predict cardiometabolic outcomes and enable clinical translation of body composition data.

## INTRODUCTION

Cross-sectional medical imaging, such as magnetic resonance imaging (MRI) and computed tomography (CT) scans, have become a fundamental tool in modern healthcare systems to support diagnosis, disease monitoring, and prognosis, with an 86% increase in MRI and a 127% increase in the volume of CT scans in England over the last decade (*1, 2*). Growing evidence shows that body composition measures, such as adipose tissue compartments and skeletal muscle derived from MRI and CT scans, are critical and independent risk factors for cardiometabolic and oncological disease and mortality (*3-5*). These measures, however, are influenced by height, differ between sexes, and change substantially with aging (*3, 4, 6-8*). Yet, comprehensive and quantifiable reference data on how body composition typically varies across the lifespan are currently missing. Therefore, tools that adjust image-derived body composition metrics for known confounders are critical for improving screening accuracy, guiding prevention strategies, and tailoring treatment decisions by creating the potential to detect whether an individual’s body composition is at risk compared to their age-matched peers. While various cutoff values and thresholds have been proposed, they were primarily based on data from small, heterogeneous, and diseased populations (*6, 9*). The identification of high-risk body composition alterations based on age-specific reference curves could improve the comparability and generalizability of study results, thereby facilitating the translation of body composition analysis into clinical practice and personalized risk assessment. Large imaging studies of the general population, such as the UK Biobank (UKB) and the German National Cohort study (NAKO), provide this unique opportunity to establish these reference curves for body composition metrics (*10, 11*). While manual body composition measurement in large-scale imaging datasets is prohibitively time-consuming, recent advances in deep learning have enabled fully automated, accurate, and efficient quantification from cross-sectional imaging (*12, 13*).

Here, we used a fully automated deep learning framework to estimate body composition metrics, including subcutaneous adipose tissue (SAT), visceral adipose tissue (VAT), skeletal muscle (SM), intramuscular adipose tissue (IMAT), and SM fat fraction (SMFF) from whole-body MRI in over 66,000 individuals from the general population. We 1) described body composition distributions and profiles across age decades, 2) calculated age-, sex- and height-adjusted reference curves and z-scores, 3) investigated the value of these z-scores as prognostic markers to predict incident health outcomes in the general population beyond traditional clinical risk factors, and 4) made available an open-source web-based body composition z-score calculator for research and clinical use.

## RESULTS

In this study, we used a fully automated deep learning framework for comprehensive body composition analysis from whole-body MRI in a large Western European population to 1) describe age and sex specific changes across the lifespan, 2) calculate age-, sex-, and height-adjusted reference curves for these measures, 3) explored their prognostic value for cardiometabolic disease and mortality and 4) provide an open-source tool, which may help to accelerate clinical translation and comparability between research studies. An overview of the study design is provided in **Fig. 1**.

**Fig. 1.**
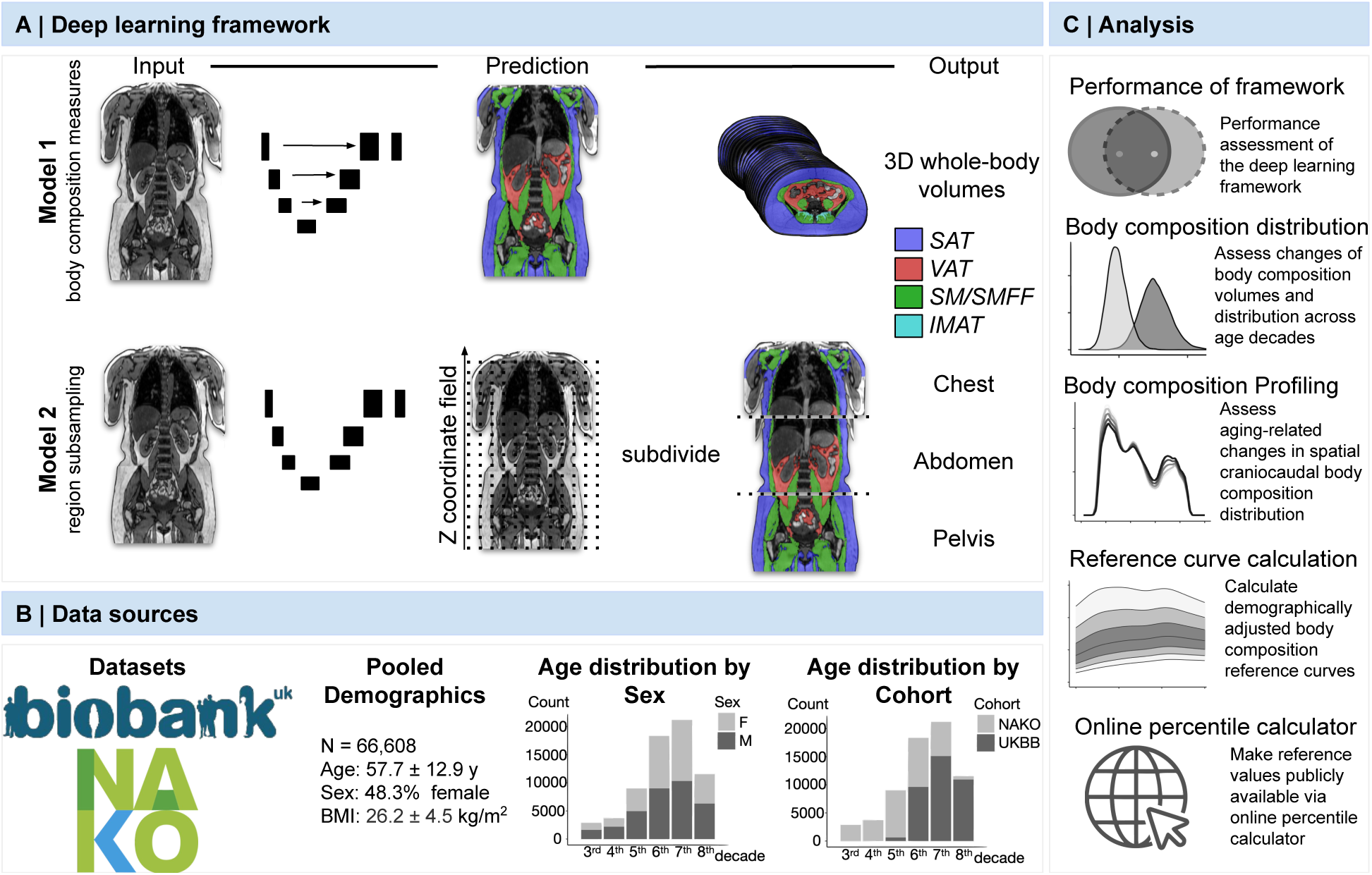
Overview of the study design. **(A)** The deep learning framework was developed to estimate body composition from MRI. The framework comprised one model (Model 1) to quantify different body composition measures (SAT, VAT, SM, SMFF, and IMAT) as 3D measures from whole-body MRI scans. A second model (Model 2) was trained to identify standardized anatomical landmarks along the craniocaudal body axis (z coordinate field), which allowed for subdividing the whole-body measures into different subregions typically examined in clinical routine MRI examination (chest, abdomen, pelvis). **(B)** Body composition was quantified from whole-body MRI in more than 66,000 individuals of two large population-based cohort studies, the UKB (36,317 individuals) and the NAKO (30,291 individuals). **(C)** After performance assessment of the framework, we investigated the change of body composition measures, distributions, and profiles across age decades, calculated age-, sex-, and height-adjusted body composition reference curves, and made them publicly available on a web-based z-score calculator. Biobank, UK Biobank. IMAT, intramuscular adipose tissue. NAKO, German National Cohort. SAT, subcutaneous adipose tissue. SM, skeletal muscle. SMFF, skeletal muscle fat fraction. VAT, visceral adipose tissue, MRI, magnetic resonance imaging

### Study population

The study population consisted of 66,608 individuals (32,165 females and 34,443 men) with a mean age of 57.7±12.9 years and a mean BMI of 26.2±4.5 kg/m^2^. In females, SAT, SMFF, and IMAT were significantly higher than in males (all p<0.001; **Table 1**), whereas males had significantly higher SM and VAT volumes (both p<0.001). Baseline demographics for the entire population and subdivided by NAKO and UKB participants are provided in **Table 1, Table S1**, and **Table S2**, respectively.

**Table 1.** Baseline characteristics - full cohort. BMI, body mass index. IMAT, intramuscular adipose tissue. L, liters SAT, subcutaneous adipose tissue. SM, skeletal muscle. SMFF, skeletal muscle fat fraction. VAT, visceral adipose tissue. Y, years

| Characteristic | Entire cohort,<br>N=66,608<sup>/</sup> | Female,<br>N=32,165<sup>/</sup> | Male,<br>N=34,443<sup>/</sup> |
| --- | --- | --- | --- |
| Age (y) | 57.7±12.9 | 58.1±12.3 | 57.3±13.4 |
| Weight (kg) | 77.3±15.8 | 69.3±13.7 | 84.7±13.9 |
| Size (m) | 1.72±0.10 | 1.64±0.07 | 1.78±0.07 |
| BMI (kg/m<sup>2</sup>) | 26.2±4.5 | 25.6±4.9 | 26.7±4.0 |
| SAT (L) | 15.72±6.68 | 17.87±7.06 | 13.71±5.61 |
| VAT (L) | 3.63±2.34 | 2.45±1.59 | 4.72±2.39 |
| SM (L) | 12.08±3.25 | 9.35±1.44 | 14.63±2.25 |
| SMFF (%) | 15.95±3.25 | 17.20±3.05 | 14.79±2.99 |
| IMAT (dL) | 1.39±0.66 | 1.48±0.63 | 1.32±0.68 |
| <sup>/</sup> Mean±SD |  |  |  |

### Differences in body composition measures across age decades

First, we analyzed the differences in median body composition measures and their IQR across age decades stratified by sex (**Fig. 2A** and **Table 2**).

**Fig. 2.**
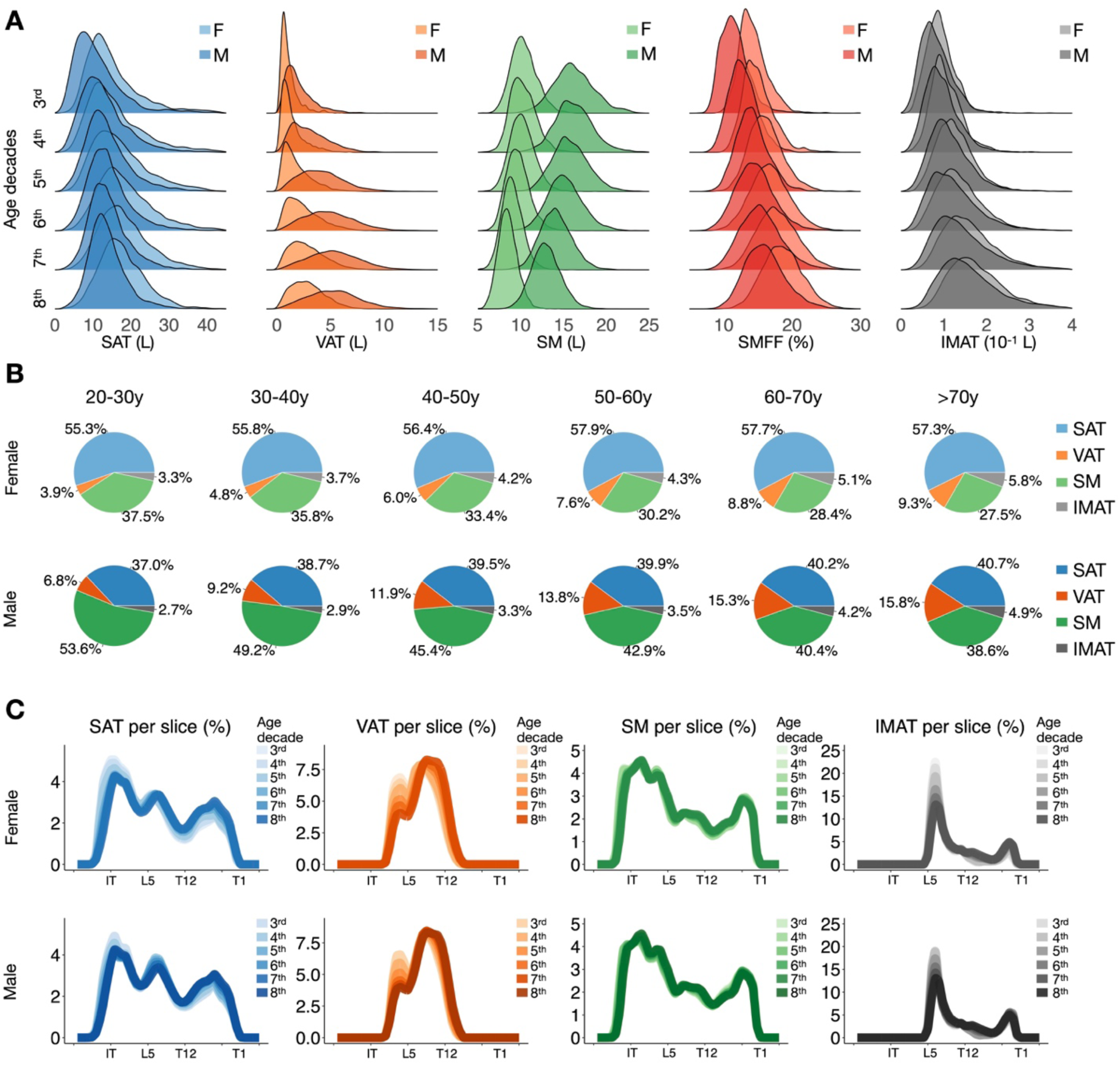
Body composition profiles across age decades. **(A)** Density plots illustrate the change in body composition measures SAT (blue), VAT (orange), SM (green), SMFF (red), and IMAT (grey) across age decades. While there is an increase in adipose tissue volume (SAT, VAT, IMAT) across age decades, there is a decrease in SM accompanied by an increase in SMFF. Median and IQR are provided in **Table 2**. **(B)** Pie charts show the age-related differences in the proportion of each body composition measure relative to the sum of all body composition measures (SAT, VAT, SM, and IMAT) for females in the top row and males in the bottom row separately. While SAT (blue) is the predominant body composition compartment in females across all age decades, it is SM (green) in males until the age of 60 years. In both females and males, SAT (blue) remains relatively stable over time, whereas there is a loss of SM (green) accompanied by a gain of VAT (orange), which is more pronounced in males than in females, and a gain of IMAT (grey), which is more pronounced in females than in males. **(C)** Profile plots demonstrate difference in the spatial distribution of each body composition measure along the craniocaudal body axis across age decades (color-coded) stratified for females (top row) and males (bottom row). During aging, SAT shifts from the gluteal region to the chest, VAT shifts from the pelvis to the abdomen, SM shifts from the gluteal region to the trunk and chest in females, and from the chest to the gluteal region in males. Paraspinal IMAT shifts from the lower lumbar spine to the upper thoracic spine. X-axis shows 50 equidistant sampling points along the craniocaudal body axis; exemplary anatomical landmarks: 10, ischial tuberosity; 20, lumbar vertebra 5; 30, thoracic vertebra 12; 43, thoracic vertebra 1. F, female. IMAT, intramuscular adipose tissue. IQR, interquartile range. L, liters. M, male. SAT, subcutaneous adipose tissue. SM, skeletal muscle. SMFF, skeletal muscle fat fraction. VAT, visceral adipose tissue. Y, years.

**Table 2.** Body composition measures (median and IQR) across age decades. IMAT, intramuscular adipose tissue. IQR, interquartile range. L, liters. SAT, subcutaneous adipose tissue. SM, skeletal muscle. SMFF, skeletal muscle fat fraction. VAT, visceral adipose tissue.

| Sex | Age | N | SAT in L | VAT in L (IQR) | SM in L | SMFF in % | IMAT in dL |
| --- | --- | --- | --- | --- | --- | --- | --- |
|  |  |  | (IQR) |  | (IQR) | (IQR) | (IQR) |
| F | 20-30y | 1238 | 13.4(10.4-18.1) | 0.85(0.6-1.3) | 10.2(9.3-11.2) | 14.3(12.8-16.1) | 0.88(0.7-1.1) |
| F | 30-40y | 1499 | 14.1(10.6-19.5) | 1.03(0.6-1.7) | 10.1(9.2-11.2) | 15.3(13.6-17.3) | 1.0(0.8-1.3) |
| F | 40-50y | 4031 | 15.7(11.8-21) | 1.46(0.8-2.5) | 10.1(9.2-11.1) | 17.0(15.1-19.3) | 1.27(1-1.6) |
| F | 50-60y | 9323 | 17.1(13.3-22.4) | 2.11(1.2-3.3) | 9.55(8.7-10.5) | 18.7(16.6-21) | 1.76(1.4-2.2) |
| F | 60-70y | 10866 | 17.3(13.5-21.9) | 2.55(1.6-3.8) | 8.91(8.1-9.8) | 20.2(18.1-22.7) | 2.16(1.8-2.6) |
| F | >70y | 5208 | 16.9(13.5-21.2) | 2.70(1.7-3.8) | 8.40(7.7-9.2) | 21.4(19.3-23.7) | 2.57(2.1-3.1) |
| M | 20-30y | 1631 | 9.64(6.8-13.8) | 1.67(1.1-2.6) | 16.0(14.6-17.6) | 11.9(10.3-13.6) | 0.76(0.6-1) |
| M | 30-40y | 2198 | 11.6(8.5-15.6) | 2.74(1.6-4.1) | 16.0(14.7-17.5) | 13.6(12-15.4) | 0.89(0.7-1.2) |
| M | 40-50y | 4964 | 12.7(9.7-16.5) | 3.92(2.6-5.4) | 15.6(14.4-17.1) | 15.4(13.5-17.3) | 1.10(0.8-1.5) |
| M | 50-60y | 9014 | 13.2(10.3-16.7) | 4.70(3.2-6.3) | 15.0(13.7-16.4) | 16.4(14.5-18.6) | 1.54(1.2-2) |
| M | 60-70y | 10322 | 13.2(10.5-16.6) | 5.21(3.6-6.9) | 14.0(12.8-15.3) | 17.7(15.6-20.1) | 1.99(1.5-2.5) |
| M | >70y | 6314 | 12.9(10.4-16.1) | 5.19(3.6-6.8) | 12.9(11.8-14) | 18.7(16.7-21.1) | 2.48(2-3.1) |

We observed a positive association between SAT and age, with a minor decrease in variability across the lifespan that was more pronounced in females (**Fig. 2A**, **Table 2**). VAT increased throughout the lifespan with higher variability in older adults in both sexes, but to a much greater extent in males (**Fig. 2A**, **Table 2**). While SM began to notably decline and become less variable after age 50, SMFF and IMAT increased and became more variable across all decades (**Fig. 2A**, **Table 2**). Similar results were seen in stratified analysis of the NAKO (**Fig. S1A & Table S3**) and UKB (**Fig. S2A & Table S4**).

### Relative proportion of body composition measures across age decades

Next, we assessed the assessed the proportion of each body composition measure relative to the sum of all body composition measures (SAT, VAT, SM, and IMAT) across age decades stratified by sex (**Fig. 2B**). Across all age decades, SAT was the predominant body composition measure in females (**Fig. 2B, top row**) exhibiting only minor differences from a minimum of 55.3% at 20-30 years to a maximum of 57.3% at 50-60 years (males: 37.0% at 20-30 years to a maximum of 40.2% at 60-70 and >70 years; **Fig. 2B, bottom row**). In contrast, SM was the predominant body composition measure in males from 20 to 70 years (**Fig. 2B, bottom row**) that steadily decreased from 53.6% at 20-30 years to 38.6% at >70 years (females: 37.5% at 20-30 years to 27.5% at >60 years; **Fig. 2B, top row**). While the IMAT proportion increased slightly from 20-30 years to >70 years (females: 3.3% to 5.8%; males: 2.7% to 4.9%), the relative VAT proportion increased substantially across age decades (females: 3.9% to 9.3%; **Fig. 2B, top row**; males: 6.8% to 15.8%; **Fig. 2B, bottom row**). Similar results were seen in stratified analysis of the NAKO (**Fig. S1B**) and UKB (**Fig. S2B**).

In addition, differences in the relative proportion of body composition measures per BMI categories are presented in **Fig. S3** for the entire study cohort and the NAKO and UKB separately.

### Spatial differences in body composition distribution along the craniocaudal body axis

We then analyzed differences in the spatial distribution of body composition measures along the craniocaudal body axis across age decades stratified by sex (**Fig. 2C**).

In both females and males, SAT shifted from the gluteal region to the chest and VAT shifted from the pelvis to the abdomen with increasing age (**Fig. 2C**). These differences were less pronounced for SM, but we observed a slight shift from the gluteal region to the trunk and chest in females (**Fig. 2C**), and a slight shift from the chest to the gluteal region in males (**Fig. 2C**). Regarding paraspinal IMAT, there was a shift from the lower lumbar spine to the upper thoracic spine in both sexes (**Fig. 2C**). Similar results were found in stratified analysis of the NAKO (**Fig. S1C**) and, to a lesser extent, in the relatively older UKB (**Fig. S2C**).

### Age, sex, and height-adjusted body composition reference curves

Next, we calculated age-, sex-, and height-specific reference curves for all body composition measures (**Fig. 3**). Crude values for each body composition measure across age stratified by sex are shown as a scatter plot in **Fig. 3A**. Reference curves were calculated using a GAM fit stratified by sex using smooth functions of age and height (see **Methods**). Examples of the resulting reference curves for each body composition measure are shown for an average female (1.65 m tall; **Fig. 3B**) and an average male (1.75 m tall; **Fig. 3C**). While the variance in SAT volume was higher in average females (**Fig. 3B**), the variances of VAT and SM were higher in average males across all ages (**Fig. 3C**). Variances were comparable for SMFF and IMAT for both sexes (**Fig. 3B & 3C**).

**Fig. 3.**
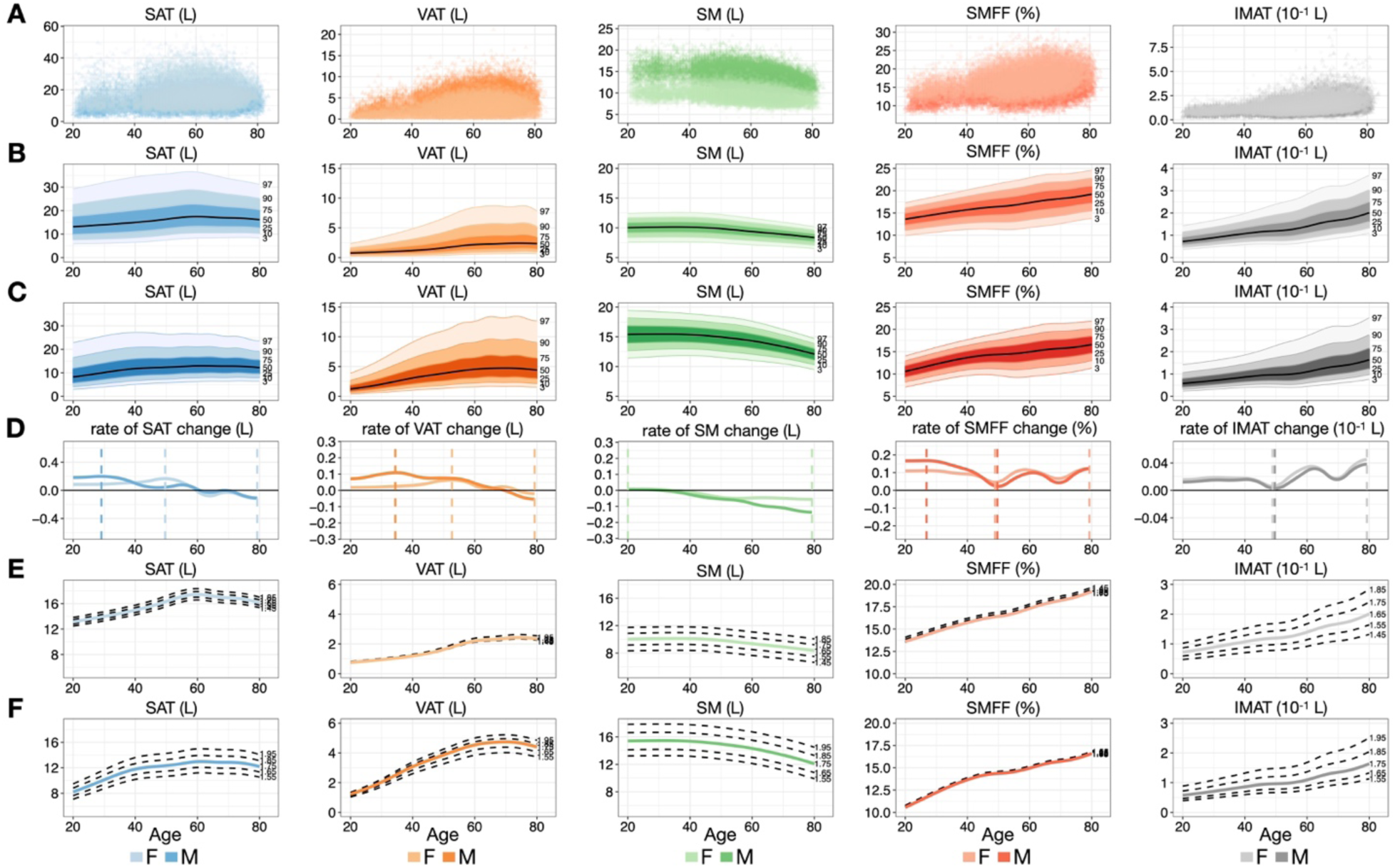
Reference curves for body composition measures. **(A)** Scatterplots of crude body composition measures as a function of age (stratified by sex). **(B & C)** Graphs show age-, sex-, and height-adjusted body composition reference curves with 3^rd^, 10^th^, 25^th^, 50^th^, 75^th^, 90^th^, and 97^th^ percentile lines for a 1.65 m tall female **(B)** and a 1.75 m tall male **(C)**. SAT variance was higher in 1.65m tall females **(B)**. Variances of VAT and SM were higher in 1.75m tall males than females across all ages (**C**). Variances were comparable for SMFF and IMAT for both sexes **(B & C)**. **(D)** The derivatives of the 50^th^ percentiles were used to illustrate representative growth curves of each body composition measure over the lifespan (stratified by sex). Dashed lines indicate the minimum and maximum rate of change. **(E & F)** Graphs show the 50^th^ percentile of females **(E)** and males **(F)** of different body heights (female: 1.45-1.85 m; male: 1.55-1.85 m). SAT and VAT volumes were less height-dependent in females (**D**) than in males (**E**). While SM and IMAT volumes were roughly similar height-dependent in males and females, DIXON-derived SMFF was nearly height-independent **(D & E)**. IMAT, intramuscular adipose tissue. L, liter. M, meter. SAT, subcutaneous adipose tissue. SM, skeletal muscle. SMFF, skeletal muscle fat fraction. VAT, visceral adipose tissue.

The derivatives of the 50^th^ percentiles were calculated to illustrate the rate of change of each body composition measure over the lifespan of an average tall female and male (**Fig. 3D)**. SAT and VAT were increasing in volume until 60 or 70 years when they started to decrease (**Fig. 3D)**. In contrast, a decline in SM occurred significantly earlier, starting at age 30 for both sexes (**Fig. 3D)**. SMFF and IMAT increased throughout the lifespan (**Fig. 3D)**. SAT and VAT were less height-dependent in females (**Fig. 3E**) than in males (**Fig. 3F**). While SM and IMAT were quite similarly height-related in males and females, SMFF showed almost no height dependency (**Fig. 3E & 3F**).

### Association between body composition z-scores and health outcomes

Last, we investigated the prognostic value of body composition z-scores for cardiometabolic outcomes in the UKB. This cohort consisted of 34,638 individuals (18,267 females and 16,371 males) with a mean age of 64.9±7.8 years and a mean BMI of 25.8±4.2 kg/m^2^ after excluding individuals with prevalent diabetes or a history of myocardial infarction and/or stroke before the date of their MRI examination (**Table S5**). Cumulative incidence and Kaplan-Meier curves showed graded associations between body composition z-score categories and outcomes (**Fig. S4**).

#### Diabetes

Over a median of 4.75 years (IQR 3.89-6.08 years), 657 individuals (1.9%) were diagnosed with diabetes. In Cox regression adjusted for age, sex, and BMI category, race, alcohol consumption, smoking status, hypertension, and history of cancer, the high z-score categories of VAT (aHR: 2.69, 95% CI [2.22-3.27]), SMFF (aHR: 1.74, 95% CI [1.45-2.09]), and IMAT (aHR: 1.22, 95% CI [1.01-1.47]), as well as the low z-score category of SM (aHR: 1.36, 95% CI [1.12-1.65]), were associated with a higher risk, while the low z-score categories of VAT (aHR: 0.54, 95% CI [0.35-0.85]) and SMFF (aHR: 0.45, 95% CI [0.30-0.68]) were associated with a lower risk of incident diabetes (**Fig. 4A**).

**Fig. 4.**
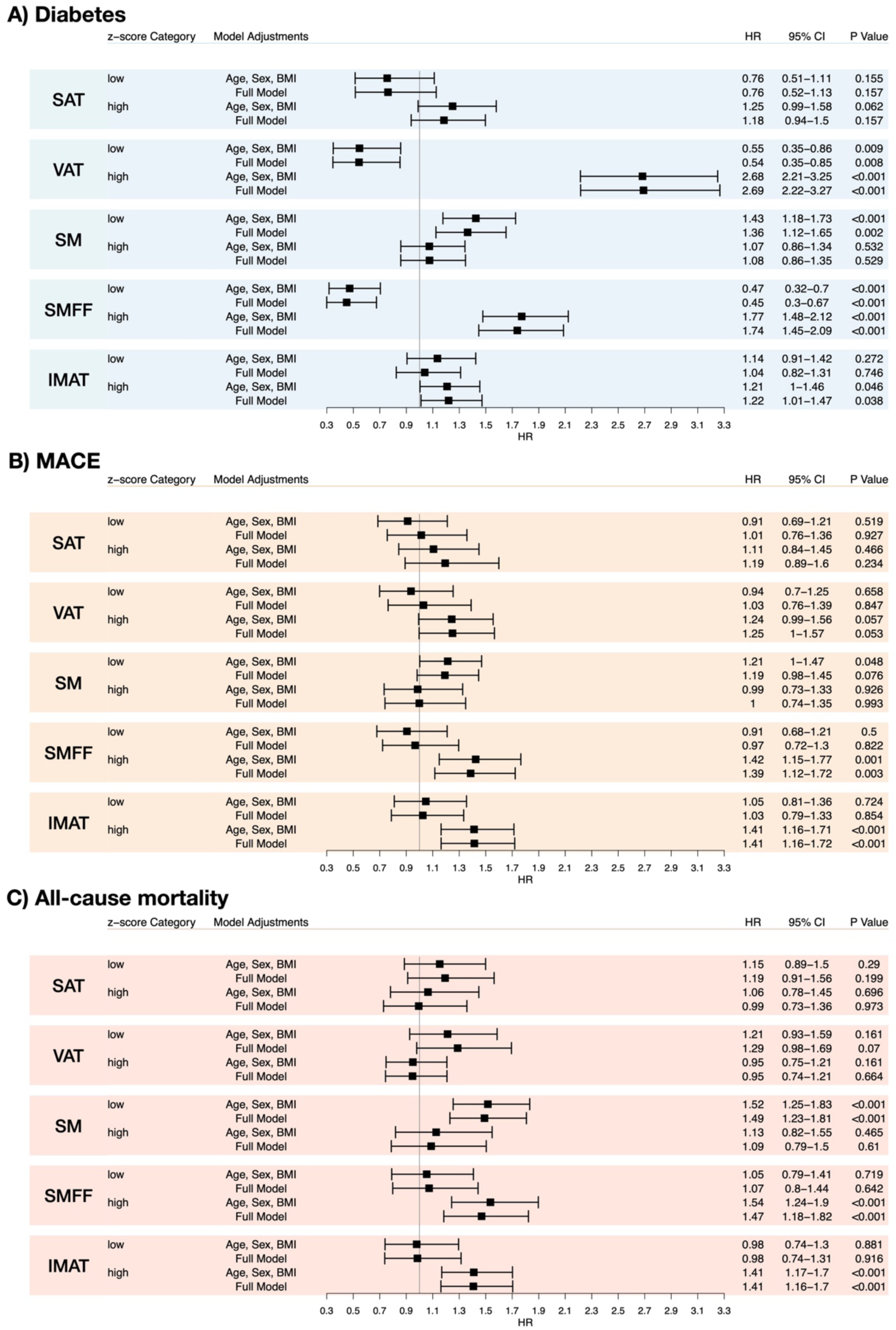
Forest plots for the body composition z-score categories and incident diabetes, MACE, and all-cause mortality in the UKB. Forest plots show hazard ratios for high and low z-score categories compared to middle categories with 95% confidence intervals of Cox proportional hazard regression analysis for (**A**) incident diabetes, (**B**), incident MACE, and (**C**) all-cause death in the UKB after excluding individuals with prevalent diabetes or a history of myocardial infarction and/or stroke (n = 34,638). Models were adjusted for 1) age, sex, and BMI category (upper row per z-score category), and 2) additional adjustment for traditional risk factors (lower row per z-score category) including race, alcohol consumption, smoking status, hypertension, and history of cancer (full model) for each body composition measure. IMAT, intramuscular adipose tissue. SAT, subcutaneous adipose tissue. SM, skeletal muscle. SMFF, skeletal muscle fat fraction. VAT, visceral adipose tissue. UKB, UK Biobank, BMI, body mass index, MACE, major adverse cardiovascular events, HR, hazard ratio, CI, confidence interval

#### MACE

Over a median of 4.76 years (IQR 3.90-6.12 years), 575 individuals (1.7%) had a MACE. In Cox regression adjusted for the above-mentioned risk factors, the high z-score category for SMFF (aHR: 1.39, 95% CI [1.12-1.72]) and IMAT (aHR: 1.41, 95% CI [1.16-1.72]) were independently associated with a higher risk of MACE (**Fig. 4B**).

#### All-cause mortality

Over a median follow-up of 4.78 years (IQR 3.93 – 6.15 years), 573 deaths (1.7%) were observed. In multivariable adjusted Cox regression, the low z-score category of SM (aHR: 1.49, 95% CI [1.23-1.81]) and the high z-score category for SMFF (aHR: 1.47, 95% CI [1.18-1.82]) and IMAT (aHR: 1.41, 95% CI [1.16-1.70]) were independent predictors of all-cause mortality (**Fig. 4C**).

## DISCUSSION

In this study, we used a fully automated deep learning framework for robust and accurate quantification of body composition measures from whole-body MRI to investigate and describe typical changes, distributions and profiles across the lifespan, and to calculate sex-stratified reference curves for adipose tissue compartments and SM in a large Western European population of more than 66,0000 individuals. Our major findings were that 1) adipose tissue compartments increase and SM decreases in both sexes throughout the lifespan, with greater variability in SAT among females and VAT and SM among males, 2) adipose tissue compartments demonstrate a cranial shift over the lifespan in both sexes; SM shifts from the gluteal region to the chest in females, while the opposite was observed in males, and 3) body composition z-score categories were independent predictors of cardiometabolic outcomes in the general population. Our study may provide the basis for tracking physiological and pathological changes in body composition across the lifespan, thereby improving personalized risk assessment and subsequent clinical decision-making. In addition, our publicly available body composition z-score calculator addresses a fundamental need in research and clinical settings: a tool that provides reference values from the general population, thereby significantly improving the comparability and generalizability of body composition measures in research and clinical practice (https://circ-ml.github.io/).

Most previous studies have used published cutoff values from small or heterogeneous patient cohorts or have calculated optimized threshold values for their study cohorts.(*14, 15*) In addition, there is no consensus on how to account for sex-, height-, and especially age-related biases.(*16-20*) To address these limitations, we released an open-source online tool along with this study to allow researchers and clinicians to compare body composition measures from their study cohort or patients against age-, sex-, and height-normalized values from a large sample of the general population. This approach has the potential to 1) improve the comparability, interpretability, reproducibility, and generalizability of research studies, and 2) translate quantitative body composition assessment to clinical practice.

One clinical implementation is an opportunistic screening strategy, where body composition data is automatically extracted from routine imaging scans, regardless of their initial indication. The online tool includes reference values and z-scores for common clinical scan regions (chest, abdomen, pelvis) and a single two-dimensional slice at the height of the L3 vertebra, which is a broadly established approach to quantify body composition.(*21, 22*) Thus, our online tool can integrate currently unused but potentially prognostic body composition data from routine imaging into the Electronic Medical Record (EMR), benchmarking it against a large Western population without disrupting established clinical workflows.

As there is increasing evidence that measures of body composition provide important prognostic information in cardiometabolic and oncologic diseases and may be related to treatment tolerability and outcomes, our reference curves may help to identify individuals with abnormal body composition measures from routine imaging examinations.(*4, 6, 8, 23*) This could be used 1) to identify prefrail individuals or 2) as a novel approach for accurate personalized dosing of systemic drug therapies including chemotherapy and immunotherapy dosing.(*7, 8, 24*) Other future applications could include tracking body composition changes from sequential imaging examinations, which may provide important physiological information on treatment tolerability or incremental prognostic information on survival from restaging imaging examinations.(*25, 26*) In addition, tracking changes in body composition may serve as a robust endpoint for the trial of lifestyle, surgical, and novel pharmacological interventions such as the recently introduced Glucagon-like peptide-1 (GLP-1) agonists for diabetes and weight loss.(*27*) In research, our framework applied to routine imaging examinations could generate large clinical data sets for further studies, to better understand the role of body composition-derived z-scores for disease-specific risks and clinical outcomes.(*28*)

This study has limitations. First, the study population is predominantly white Western European adults over the age of 20 from the UK and Germany. The generalizability of the reference curves to other racial/ethnic groups, children, and/or patients in a clinical setting may be limited. Second, despite the possibility of integrating the framework into clinical practice, whole-body MRI is not a commonly performed examination. To account for this, we also provided reference curves for typical anatomical regions covered in routine MRI (i.e., chest, abdomen, pelvis) and a single slice at the L3 vertebra, which is currently the most widely used approach to quantify body composition.

In conclusion, we automatically quantified body composition from whole-body MRI from a large Western European population to describe aging-related differences and to calculate age, sex, and height-specific reference curves for body composition measures (SAT, VAT, SM, SMFF, and IMAT). Alterations from body composition reference values were predictors of cardiometabolic outcomes in the general population beyond traditional risk factors. To support current and future research and accelerate clinical translation, we released an open-source online tool that allows researchers and clinicians to normalize their own data sets to improve comparability and generalizability of body composition research.

## MATERIAL AND METHODS

### Study design

In this study, we used a publicly available deep learning framework to quantify three-dimensional (3D) body composition automatically measures from whole-body MRI and common anatomical regions covered in routine clinical MRI examinations (i.e., chest, abdomen, pelvis) (*5*). Body composition measures included SAT, VAT, SM, SMFF, and IMAT. First, we analyzed body composition measure density plots, their relative proportions, and their distributions along the craniocaudal body axis across the lifespan. Second, we calculated age-, sex-, and height-specific z-scores for all body composition measures. Third, we investigated the association between these z-scores with incident health outcomes. Lastly, we developed an open-source web-based calculator to enable the comparison of body composition metrics against reference values from a large Western European population.

### Data sources

This study used data from two large population-based cohort studies designed to examine disease prevention and prognosis in individuals from the general population: 1) the UKB and 2) the NAKO (*11, 29*). Between 2006-2010, 500,000 individuals aged 40-69 (5.5% of invitees) joined the UKB after NHS invitation (*30*). The NAKO is ongoing, with 205,415 participants aged 19-74 years enrolled at 18 sites in Germany (*31*). In both studies, a comprehensive MRI protocol was acquired in a subgroup of participants. In this study, we used the whole-body T_1_-weighted 3D VIBE two-point DIXON technique from the UKB and NAKO to quantify body composition. Detailed information on the data sources is provided in **Supplemental Methods**. Flow charts for the data sources are provided in **Fig. S5 & S6**.

### Deep learning framework

We used a fully automated open-source deep learning framework optimized for BC quantification in the UKB and NAKO to volumetrically quantify whole-body composition metrics including SAT, VAT, SM, SMFF, and IMAT from whole-body MRI. The model exhibited high performance for BC quantification, with Dice coefficients for the different BC measures of ≥0.88 in the NAKO and ≥0.86 in the UKB testing datasets, indicating reliable results for subsequent analyses. Further detailed information on model development and testing is provided elsewhere (*5*).

### Outcomes in the UKB

All survival analyses were performed in the UKB only, as outcome data were not available for the NAKO. Incident outcomes in the UKB were 1) diabetes (ICD-10: E10-14; ICD-9: 250), 2) major adverse cardiovascular events (MACE) defined as myocardial infarction (ICD-10: I21-22; ICD-9: 410-411), ischemic stroke (ICD-10: I63; ICD-9: 433-434), or mortality from major cardiovascular diseases (ICD-10: I00-I78), and 3) all-cause mortality. Outcomes in the UKB were defined based on ICD-10 and ICD-9 codes through linkage to electronic medical records or national death registries. Follow-up time in the UKB was calculated as the date of the MRI examination until the earliest date among date of death, outcome, loss to follow-up, or May 25, 2023 (date of UKB data download).

### Covariates

The following a priori defined covariable were included in the analyses: age, sex, BMI (height/m^2^), race, smoking status (never, former, current), alcohol consumption, hypertension, and history of cancer. Further detailed information on the extraction and definition of covariates from UKB and NAKO is provided in **Supplemental Methods.**

### Statistical Analysis

#### Data harmonization

We observed a small distribution shift for SMFF and IMAT between the UKB and NAKO. To account for this, we used a normal score transformation before further analysis as detailed in **Supplemental Methods.**

#### Baseline demographics

Baseline characteristics of the study participants are presented as mean ± standard deviation (SD) or median with interquartile ranges (IQR) for continuous variables and absolute counts with percentages for categorical variables.

#### Differences in body composition across age decades

Body composition differences across age decades were visualized using density plots and quantified using the median and interquartile range (IQR) of each body composition measure per age decade. The proportion of each body composition measure relative to the sum of all body composition measures (SAT, VAT, SM, and IMAT) was presented as pie charts per age decade. In addition, the difference in the spatial distribution of body composition measures in the craniocaudal direction across age decades was described. This was done by calculating the proportion of the total volume for each body composition metric at 50 equidistant components along an axis from the femoral insertion of the adductor brevis muscle to the 1st thoracic vertebra.

#### Body composition reference curves

Finally, sex-stratified reference curves (similar to growth chart curves in pediatrics) based on age and height were created for each body composition measure for both whole-body MRI and body regions routinely covered by clinical MRI examinations (pelvis, abdomen, chest). We fit a sex-stratified generalized additive model (GAM) with isotropic smooth terms for age and height to predict the conditional mean body composition measure using the mgcv R package (version 1.9-1, 2023, open source). An additional GAM was developed for each measure using the same structure to predict the conditional variance of the body composition measure. Before modeling, log transformations were applied to adipose tissue measures (SAT, VAT, and IMAT) to mitigate skewed distributions. We performed internal 5-fold cross-validation to assess the median absolute deviation of the body composition GAMs (**Table S6**).

#### Association between age, sex, and height-specific z-scores and outcomes in the UKB

Outcome analyses were only performed in the UKB and limited to individuals without prevalent diabetes, a history of myocardial infarction, or ischemic stroke. An individual’s z-score for each body composition measure was calculated as:

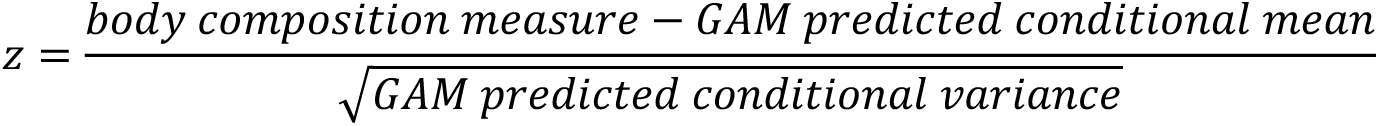

The association between age, sex, and height-specific z-scores of SAT, VAT, SM, SMFF, and IMAT and health outcomes was explored for z-score categories defined as “high” (z-score > 1), “middle” (z-score -1 to ≤ 1) and “low” (z-score < -1) for each body composition measure. Thus, an individual’s body composition measure was classified as “high” if it was greater than one standard deviation above the age-, sex-, and height-specific conditional mean. To investigate time to outcome in the UKB, cumulative incidence curves (for incident diabetes and MACE) or Kaplan-Meier survival estimates (for all-cause mortality) and log-rank tests were computed using the above-defined z-score categories. The associations between the z-score categories and health outcomes were evaluated via univariable and multivariable Cox proportional hazard regression analysis adjusted for age, sex, BMI category, race, alcohol consumption, smoking status, hypertension, and history of cancer. Results from Cox regressions were reported as hazard ratio (HR) and 95% confidence intervals (CI). All statistical analyses were performed using R V4.2.2 (R Core Team, www.r-project.org, 2022).

## Data Availability

Due to the restrictions imposed by the NAKO ethics committee, the data are not publicly available but can be requested from https://transfer.nako.de/transfer/index. The data from the UKB can be downloaded upon request from https://www.ukbiobank.ac.uk. The open-source online tool for z-score and percentile calculation is available from https://circ-ml.github.io/.

## Acknowledgments

This project was conducted with data from the German National Cohort (NAKO) (www.nako.de). The NAKO is funded by the Federal Ministry of Education and Research (BMBF) [project funding reference numbers: 01ER1301A/B/C, 01ER1511D, and 01ER1801A/B/C/D], federal states of Germany, and the Helmholtz Association, the participating universities and the institutes of the Leibniz Association. This research has been conducted using the UK Biobank Resource under Application Number 80337. We thank all participants who took part in the NAKO and UKB study and the staff of these research initiatives. MJ was funded by the Deutsche Forschungsgemeinschaft (DFG, German Research Foundation) - 518480401. VKR was funded by Norm Group Longevity Impetus Grant, NHLBI K01HL168231, and AHA Career Development Award 935176. We thank Kayden J. Kehe for developing the open-source online tool. Informed consent was obtained from all participants in the UK Biobank and the German National Cohort study. In addition, we received local IRB approval (IRB of the University of Freiburg: 23-1316-S1-retro and 24-1099-S1-retro).

## Author Contributions

Conceptualization: MJ, MR, SR, TP, TN, HUK, HV, RB, MFR, CLS, MTL, FB, VKR, JW

Data curation: MJ, MR, HR, SR, TH, TP, TN, HUK, HV, RB, MFR, CLS, MTL, FB, VKR, JW

Formal analysis: MJ, VKR, JW

Funding acquisition: SR, TP, TN, HUK, HV, RB, CLS, FB

Investigation: MJ, MR, SR, MTL, VKR, JW

Methodology: MJ, MR, HR, SR, TH, TP, TN, HUK, HV, RB, MFR, CLS, MTL, FB, VKR, JW

Project administration: MJ, SR, TP, TN, HUK, HV, RB, MFR, CLS, MTL, FB, VKR, JW

Resources: MJ, MR, HR, SR, TH, TP, TN, HUK, HV, RB, MFR, CLS, MTL, FB, VKR, JW

Software: MJ, MR, HR, SR, MFR, VKR, JW

Supervision: MR, SR, TH, TP, TN, HUK, HV, RB, MFR, CLS, MTL, FB

Visualization: MJ, VKR, JW

Writing – original draft: MJ, VKR, JW

Writing – review & editing: MJ, MR, HR, SR, TH, TP, TN, HUK, HV, RB, MFR, CLS, MTL, FB, VKR, JW

## Competing interests

Deutsche Forschungsgemeinschaft (DFG, German Research Foundation) – 518480401 (MJ)

Siemens Healthineers (Institutional support) (CLS, FB, JW)

Bayer Health (Institutional support) (CLS, FB, JW)

American Heart Association (Institutional support) (MTL)

AstraZeneca (Institutional support) (MTL)

Ionis (Institutional support) (MTL)

Johnson & Johnson Innovation (Institutional support) (MTL)

Kowa (Institutional support) (MTL)

MedImmune (Institutional support) (MTL)

National Institutes of Health / National Heart, Lung, and Blood Institute (NIH/NHLBI) (Institutional support) (MTL)

NHLBI Grant K01HL168231 (VKR)

Risk Management Foundation of the Harvard Medical Institutions Incorporated (Institutional support) (MTL)

Norn Group Longevity Impetus Grant (VKR)

American Heart Association Career Development Award 935176 (VKR)

Deutsche Forschungsgemeinschaft (DFG, German Research Foundation) – 525002713 (JW)

National Academy of Medicine, Healthy Longevity Grand Challenge Catalyst Award (MJ, VKR, JW)

## Data and materials availability

Due to the restrictions imposed by the NAKO ethics cmmittee, the data are not publicly available but can be requested from https://transfer.nako.de/transfer/index. The data from the UKB can be downloaded upon request from https://www.ukbiobank.ac.uk. The open-source online tool for z-score and percentile calculation is available from https://circ-ml.github.io/.

