## Supplemental Materials for "Aging-related changes in body composition across the lifespan – insights from over 66.000 individuals of a Western European population"

#### I MATERIAL AND METHODS

**Data sources:**

*UKB*

The UKB is a large, prospective population-based study established to investigate the genetic and non-genetic determinants of diseases of middle and older age. It included over 500,000 volunteers aged 38–73 years at baseline across the United Kingdom and provides detailed clinical parameters on prevalent diseases and outcomes (*11, 32*).

In this study, we included all UKB participants from the multimodal-imaging cohort who underwent $T_1$-weighted 3D two-point VIBE Dixon MRI in axial orientation (1.5T MAGNETOM Aera, Siemens Healthineers, Erlangen, Germany) until the date of data download, May 25, 2023 (*31*). A total of 36,515 baseline MRIs were identified. 198 individuals were excluded due to corrupt or incomplete images, resulting in a final study cohort of 36,317 individuals with available whole-body MRIs for body composition quantification (**Fig. S5**).

*NAKO*

The NAKO is an ongoing interdisciplinary epidemiologic cohort study investigating disease prevention and prognosis, focusing on major disease groups such as cardiovascular disease, diabetes, and cancer, with 205,415 participants aged 19-74 years enrolled at 18 sites in Germany (*3*). Of these, 30,861 participants underwent whole-body MRI at five imaging sites in the imaging substudy using a dedicated $T_1$-weighted 3D two-point VIBE Dixon sequence in axial orientation (3T MAGNETOM Skyra, Siemens Healthineers, Erlangen, Germany). The NAKO was approved by all local institutional review boards of the five imaging sites, and written informed consent was obtained from all participants before enrollment (*33*).

For the current study, we used MR images and clinical information from the second release that included 30,770 participants who underwent MRI between May 27, 2014 and September 30, 2019. A total of 479 individuals were excluded because of corrupt or incomplete images, resulting in a final study cohort of 30,291 individuals with available whole-body MRIs for body composition quantification (**Fig. S6**).

**Covariates:**

*UKB*

Date of birth, sex, and race were extracted from the self-reported UKB baseline population characteristics. Due to the small number of non-white participants in the UKB imaging study, race was dichotomized into white and others. Physical measurements, including BMI (weight/height$^2$; kg/m$^2$), were obtained at the imaging visit. BMI categories were defined as < 25, 25-30, and ≥ 30. Alcohol intake, smoking status, and history of cancer were self-reported via a touchscreen questionnaire at the imaging visit. Alcohol intake was defined as "regular" ("Daily or almost daily", "Three or four times a week", "Once or twice a week") and "occasional" ("One to three times a month", "Special occasions only", "Never"). Smoking was dichotomized by subsuming former and current smokers into "ever smokers". Prevalent hypertension was defined as ICD-10 codes I10-15 or ICD-9 codes 401-405.

*NAKO*

Only baseline demographic data were available for the NAKO participants. This included age at MRI, sex, height (m), and weight (kg), which were assessed with standardized measuring devices at the imaging centers (all Stadiometer 274 for height and medical Body Composition Analyzer 515 for weight, both seca GmbH, Hamburg, Germany).

**Data harmonization:**

We observed a small data shift between the UKB and NAKO data for SMFF and IMAT after extracting the body composition measures from the whole-body MRI scans.

*SMFF:* Swapping artifacts are typical artifacts in Dixon MRI caused by errors in calculating water and fat images, resulting in areas incorrectly labeled as “water” in the fat image and vice versa. This results in incorrect estimates when calculating the fat fraction extracted from the water and fat images as described above. In the UKB, swapping artifacts mainly occurred as block-wise artifacts (e.g., abdomen). These artifacts are easy to detect and correct. Therefore, all swapping artifacts in the UKB were corrected before extracting SMFF (see **Supplemental Methods** above**)**. In contrast, in the NAKO, artifacts affected only a small portion of some image slices that are difficult to detect and correct, which is likely the reason for a small right-sided distribution shift in the NAKO SMFF compared to the corrected UKB data. Since reliable correction of these small swapping artifacts is difficult, no correction algorithm has been applied. Instead, we used a normal score transformation to convert the NAKO SMFF distribution to the same mean and variance as the UKB SMFF, since the SMFF distributions were approximately normal.

*IMAT:* For IMAT, we observed a small right-shifted distribution shift in the UKB IMAT histograms compared to the NAKO IMAT histograms, likely due to the lower spatial resolution of the 1.5T VIBE Dixon sequence in the UKB. The IMAT data were harmonized by transforming the log IMAT distribution in the UKB to match that of the NAKO.

#### II SUPPLEMENTAL FIGURES

##### Fig. S1: Body composition profiles across age decades - NAKO

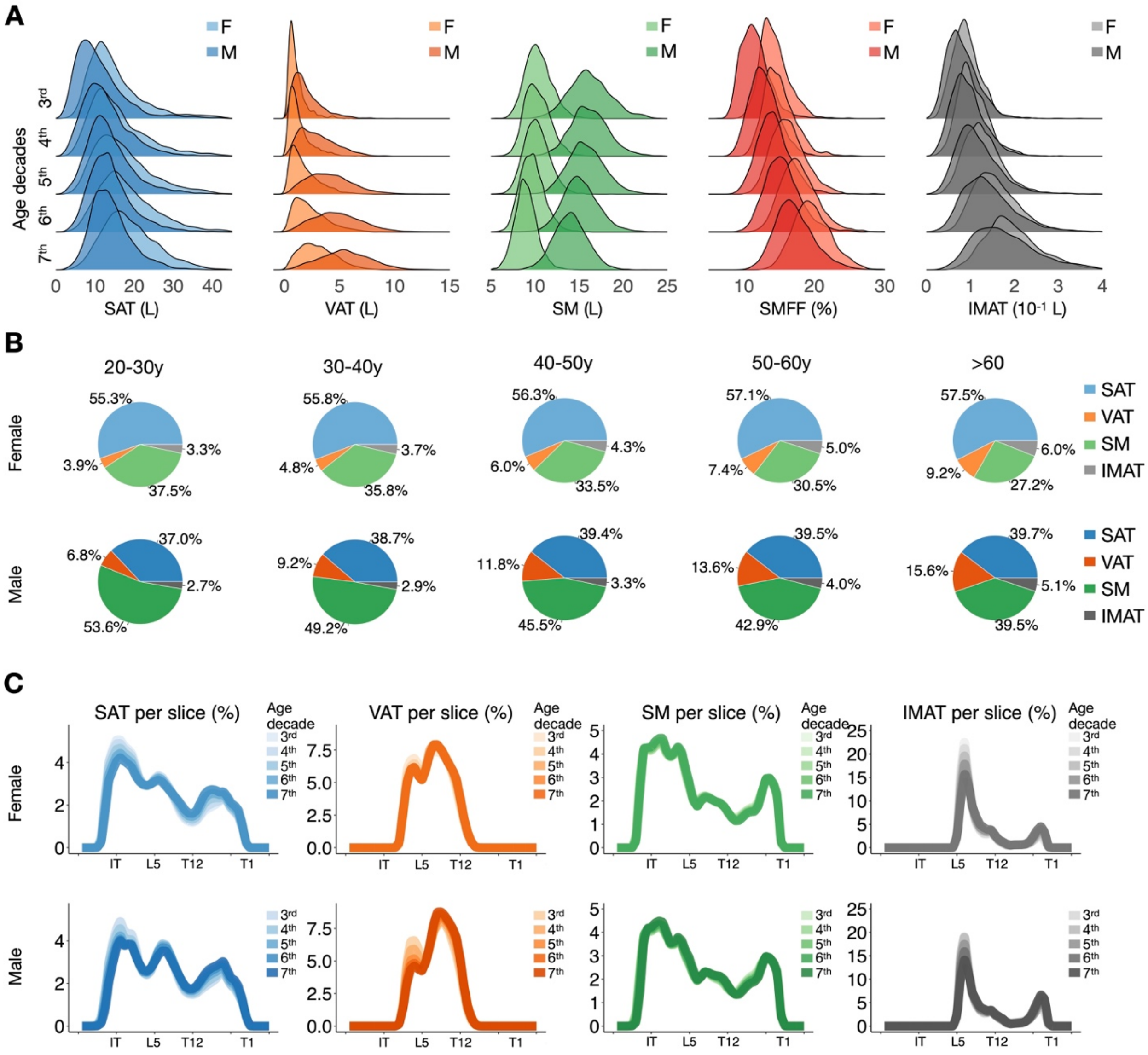

**Fig. S1 | (A)** Density plots illustrate the change in body composition measures SAT (blue)**,** VAT (orange), SM (green), SMFF (red), and IMAT (grey) across age decades in the NAKO. Median and IQR are provided in **Supplemental Table 3**. **(B)** Pie charts show the age-related differences in the proportion of each body composition measure relative to the sum of all body composition measures (SAT, VAT, SM, and IMAT) for female NAKO participants in the top row and male NAKO participants in the bottom row

separately. **(C)** Profile plots demonstrate changes in the spatial distribution of each body composition measure along the craniocaudal body axis across age decades (color-coded) stratified for females (top row) and males (bottom row) of the NAKO.
X-axis shows 50 equidistant sampling points along the craniocaudal body axis; exemplary anatomical landmarks: 10, ischial tuberosity; 20, lumbar vertebra 5; 30, thoracic vertebra 12; 43, thoracic vertebra 1.

F, female. IMAT, intramuscular adipose tissue. IQR, interquartile range. L, liters. M, male. NAKO, German National Cohort. SAT, subcutaneous adipose tissue. SM, skeletal muscle. SMFF, skeletal muscle fat fraction. VAT, visceral adipose tissue. Y, years.

**Fig. S2: Body composition profiles across age decades - UKB**

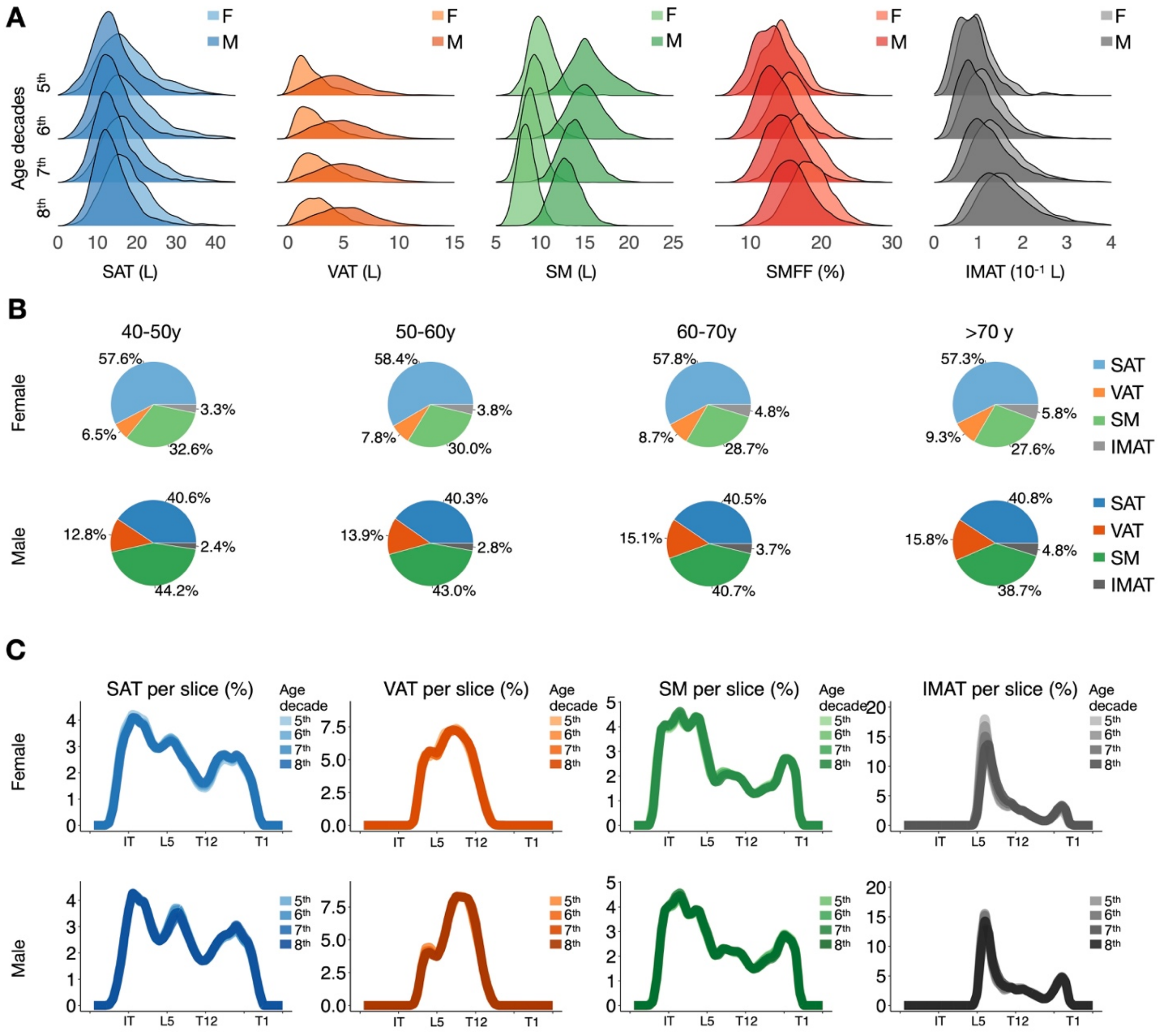

**Fig. S2 | (A)** Density plots illustrate the change in body composition measures SAT (blue), VAT (orange), SM (green), SMFF (red), and IMAT (grey) across age decades in the UKB. Median and IQR are provided in **Supplemental Table 4**. **(B)** Pie charts show the age-related differences in the proportion of each body composition measure relative to the sum of all body composition measures (SAT, VAT, SM, and IMAT) for female UKB participants in the top row and male UKB participants in the bottom row separately. **(C)** Profile plots demonstrate changes in the spatial distribution of each body composition measure along the craniocaudal body axis across age decades (color-coded) stratified for females (top row) and males (bottom row) of the UKB.

X-axis shows 50 equidistant sampling points along the craniocaudal body axis; exemplary anatomical landmarks: 10, ischial tuberosity; 20, lumbar vertebra 5; 30, thoracic vertebra 12; 43, thoracic vertebra 1.

F, female. IMAT, intramuscular adipose tissue. IQR, interquartile range. L, liters. M, male. SAT, subcutaneous adipose tissue. SM, skeletal muscle. SMFF, skeletal muscle fat fraction. UKB, UK Biobank. VAT, visceral adipose tissue. Y, years.

**Fig. S3: Relative proportion of each body composition measure by BMI categories**

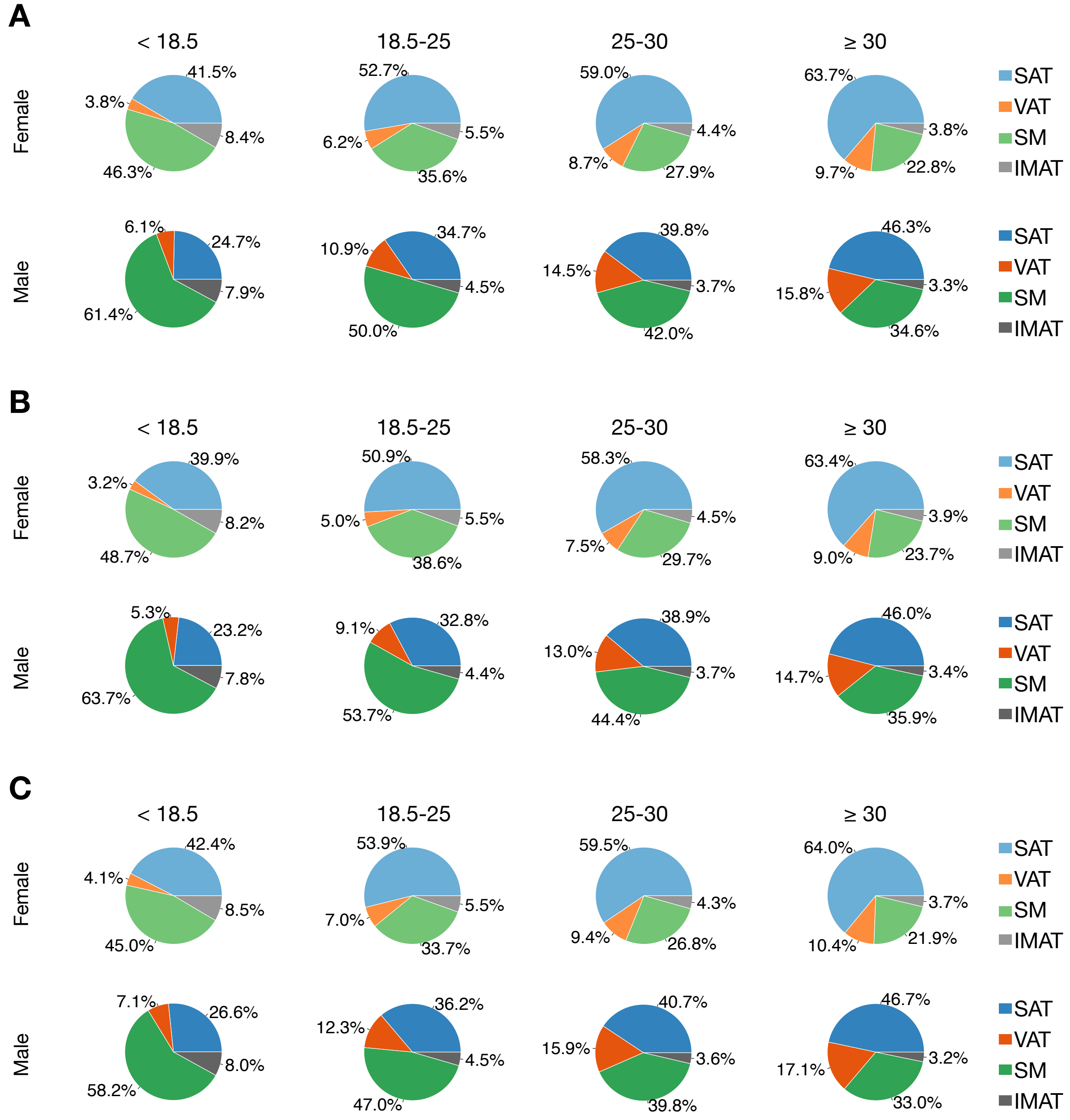

**Fig. S3 |** Pie charts show the change of relative percentage of each body composition compartment to the sum of the body composition measures (SAT, VAT, SM, and IMAT) across BMI categories stratified by sex for the entire cohort **(A)**, the NAKO **(B)**, and the UKB **(C)**.

BMI, body mass index. IMAT, intramuscular adipose tissue. NAKO, German National Cohort. SAT, subcutaneous adipose tissue. SM, skeletal muscle. SMFF, skeletal muscle fat fraction. UKB, UK Biobank. VAT, visceral adipose tissue.

#### Fig. S4: Cumulative incidence and Kaplan-Meier curves for outcomes in the UKB

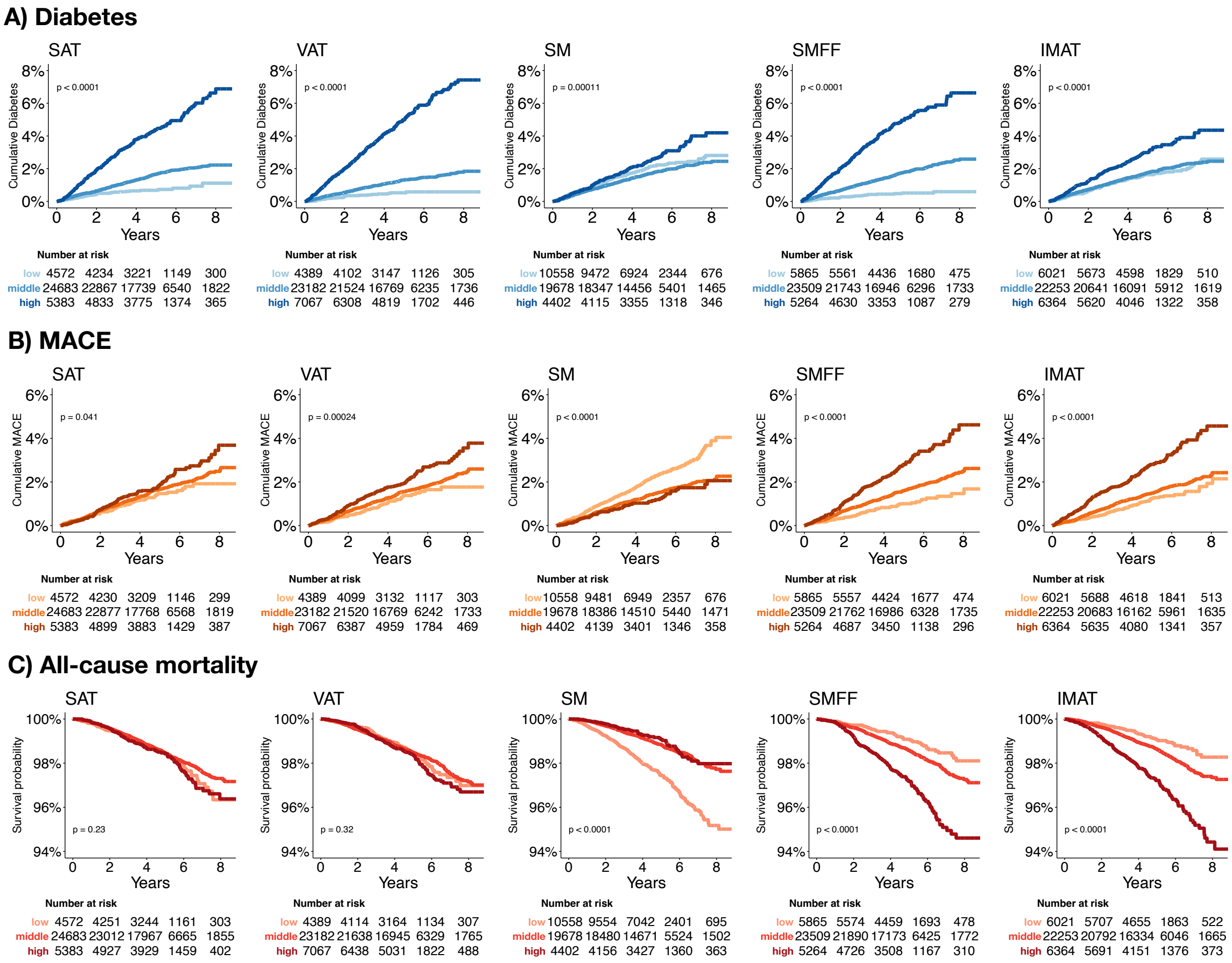

**Fig. S4 |** Cumulative incidence curves for (**A**) Diabetes and (**B**) MACE, and Kaplan Meier curves for (**C**) all-cause mortality according to low (z<-1), middle (-1≤z≤1) and high (z>1) z-score categories in the UKB. Cumulative incidence curves (**A & B**) show significant differences between all body composition measure z-score categories. Kaplan Meier plots (**C**) show significant differences between SM, SMFF, and IMAT z-score categories.

IMAT, intramuscular adipose tissue. MACE, major adverse cardiovascular event. SAT, subcutaneous adipose tissue. SM, skeletal muscle. SMFF, skeletal muscle fat fraction. UKB, UK biobank. VAT, visceral adipose tissue.

**Fig. S5: Flowchart Diagram-UKB**

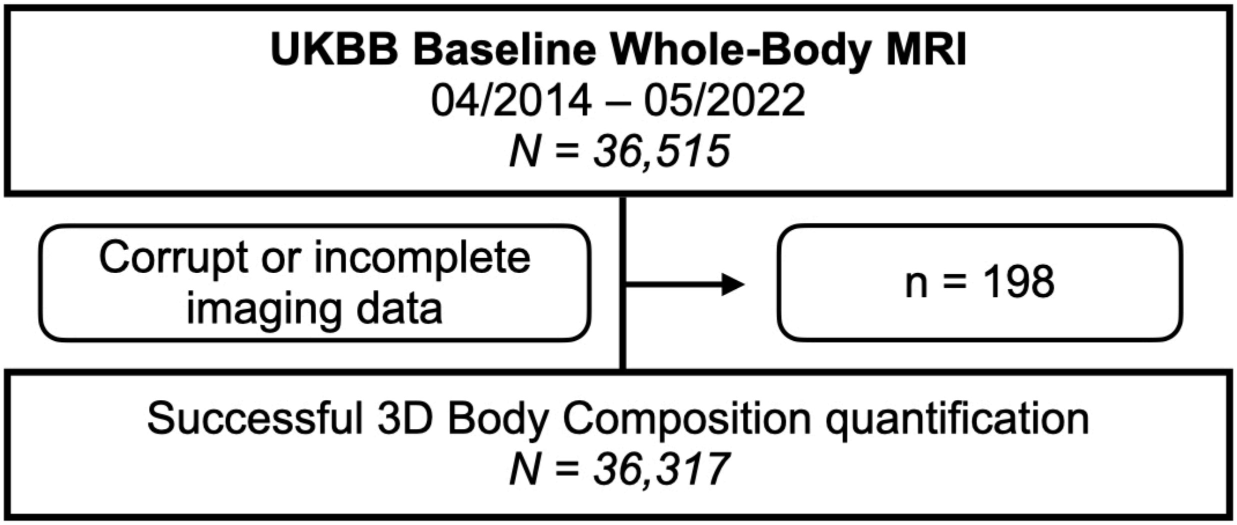

Body composition was defined as subcutaneous adipose tissue, visceral adipose tissue, skeletal muscle, and intramuscular adipose tissue.

MRI, magnetic resonance imaging. UKB, UK Biobank

**Fig. S6: Flowchart Diagram-NAKO**

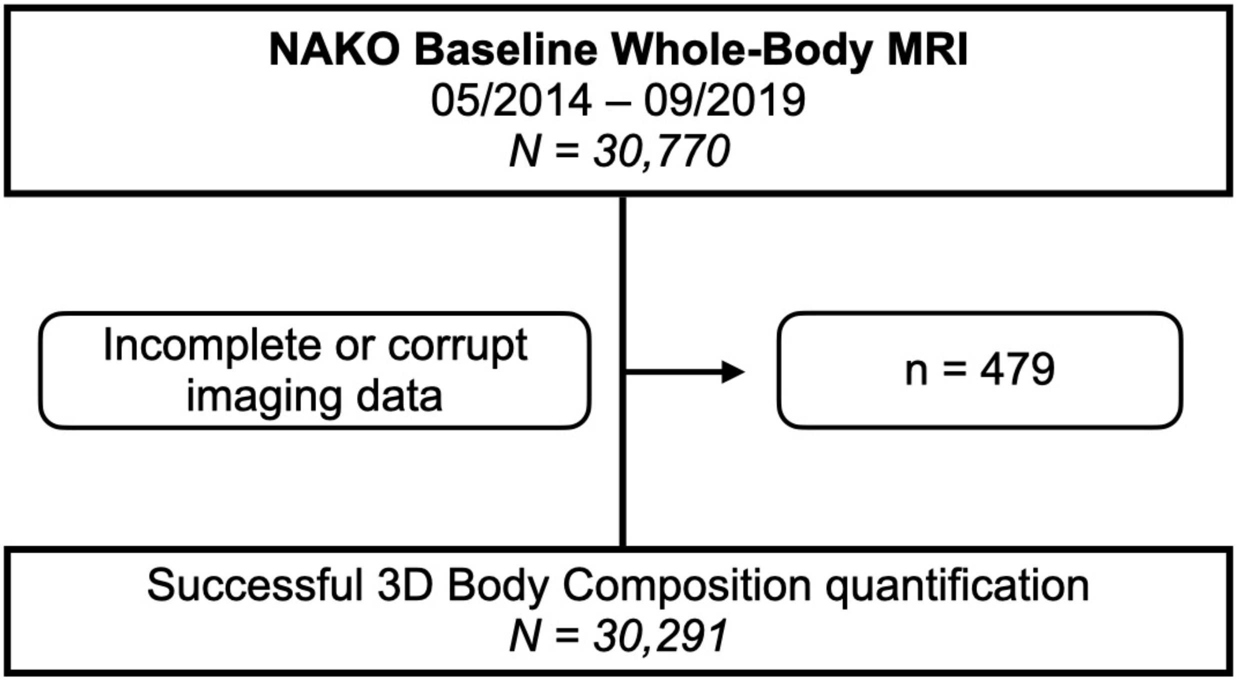

Body composition was defined as subcutaneous adipose tissue, visceral adipose tissue, skeletal muscle, and intramuscular adipose tissue.

MRI, magnetic resonance imaging. NAKO, German National Cohort.

### III SUPPLEMENTAL TABLES

##### Table S1: Baseline demographics - NAKO

| Characteristic | **Overall**, N=30,291[1] | **Female**, N=13,388[1] | **Male**, N=16,903[1] |
|---|---|---|---|
| Age (y) | 48.9±12.3 | 49.3±12.2 | 48.6±12.4 |
| Weight (kg) | 79.8±16.4 | 71.2±14.6 | 86.6±14.3 |
| Height (m) | 1.73±0.10 | 1.66±0.07 | 1.79±0.07 |
| BMI (kg/m$^2$) | 26.5±4.7 | 26.0±5.3 | 27.0±4.2 |
| SAT (L) | 15.25±6.85 | 17.50±7.38 | 13.47±5.82 |
| VAT (L) | 3.32±2.25 | 2.13±1.51 | 4.27±2.29 |
| SM (L) | 12.86±3.36 | 9.76±1.48 | 15.31±2.22 |
| SMFF (%) | 15.86±3.25 | 17.20±3.05 | 14.79±2.99 |
| IMAT (dL) | 1.39±0.65 | 1.48±0.61 | 1.32±0.67 |

[1] Mean±SD

BMI, body mass index. IMAT, intramuscular adipose tissue. L, liters SAT, subcutaneous adipose tissue. SM, skeletal muscle. SMFF, skeletal muscle fat fraction. VAT, visceral adipose tissue. Y, years

**Table S2: Baseline demographics - UKB**

| **Characteristic** | **Overall**, N=36,317[1] | **Female**, N=18,777[1] | **Male**, N=17,540[1] |
|---|---|---|---|
| Age (y) | 65.1±7.8 | 64.4±7.7 | 65.7±7.9 |
| Weight (kg) | 75.2±15.0 | 68.0±12.8 | 83.0±13.2 |
| Height (m) | 1.70±0.09 | 1.64±0.06 | 1.77±0.07 |
| BMI (kg/m2) | 25.9±4.3 | 25.4±4.6 | 26.4±3.8 |
| SAT (L) | 16.11±6.51 | 18.14±6.81 | 13.94±5.39 |
| VAT (L) | 3.88±2.38 | 2.68±1.60 | 5.16±2.40 |
| SM (L) | 11.43±3.01 | 9.06±1.33 | 13.97±2.08 |
| SMFF (%) | 16.04±3.25 | 17.20±3.05 | 14.79±2.99 |
| IMAT (dL) | 1.40±0.67 | 1.48±0.63 | 1.32±0.69 |

[1] Mean±SD

BMI, body mass index. IMAT, intramuscular adipose tissue. L, liters SAT, subcutaneous adipose tissue. SM, skeletal muscle. SMFF, skeletal muscle fat fraction. VAT, visceral adipose tissue. Y, years

**Table S3: Body composition measures (median and IQR) across age decades - NAKO**

| Sex | Age | N | SAT in L (IQR) | VAT in L (IQR) | SM in L (IQR) | SMFF in % (IQR) | IMAT in dL (IQR) |
|---|---|---|---|---|---|---|---|
| F | 20-30y | 1238 | 13.4(10.4-18.1) | 0.85(0.6-1.3) | 10.2(9.3-11.2) | 14.3(12.8-16.1) | 0.88(0.7-1.1) |
| F | 30-40y | 1499 | 14.1(10.6-19.5) | 1.03(0.6-1.7) | 10.1(9.2-11.2) | 15.3(13.6-17.3) | 1.0(0.8-1.3) |
| F | 40-50y | 3675 | 15.7(11.7-21) | 1.45(0.8-2.5) | 10.1(9.2-11.1) | 17.1(15.1-19.3) | 1.24(1-1.5) |
| F | 50-60y | 3941 | 16.8(12.9-22.1) | 2.02(1.2-3.2) | 9.57(8.7-10.5) | 19.0(16.9-21.3) | 1.51(1.2-1.9) |
| F | >60y | 3035 | 17.9(14-22.9) | 2.84(1.8-4.1) | 8.88(8.1-9.7) | 21.5(19.3-24.1) | 1.88(1.5-2.3) |
| M | 20-30y | 1631 | 9.64(6.8-13.8) | 1.67(1.1-2.6) | 16.0(14.6-17.6) | 11.9(10.3-13.6) | 0.76(0.6-1) |
| M | 30-40y | 2198 | 11.6(8.5-15.6) | 2.74(1.6-4.1) | 16.0(14.7-17.5) | 13.6(12-15.4) | 0.89(0.7-1.2) |
| M | 40-50y | 4682 | 12.6(9.7-16.5) | 3.90(2.6-5.4) | 15.7(14.4-17.1) | 15.4(13.6-17.3) | 1.07(0.8-1.4) |
| M | 50-60y | 4776 | 13.1(10.1-16.6) | 4.66(3.2-6.2) | 15.0(13.8-16.4) | 16.9(15-19) | 1.33(1-1.7) |
| M | >60y | 3616 | 13.2(10.4-16.7) | 5.45(4-7) | 14.0(12.7-15.2) | 19.0(16.9-21.5) | 1.69(1.3-2.2) |

IMAT, intramuscular adipose tissue. IQR, interquartile range. L, liters. SAT, subcutaneous adipose tissue. SM, skeletal muscle. SMFF, skeletal muscle fat fraction. VAT, visceral adipose tissue.

**Table S4: Age-related changes in body composition measures (median and IQR) - UKB**

| Sex | Age | N | SAT in L (IQR) | VAT in L (IQR) | SM in L (IQR) | SMFF in % (IQR) | IMAT in dL (IQR) |
|---|---|---|---|---|---|---|---|
| F | 40-50y | 356 | 16.5(12.7-21.6) | 1.65(1-2.8) | 10.0(9.1-11) | 16.9(15.2-19.1) | 1.69(1.3-2) |
| F | 50-60y | 5382 | 17.5(13.6-22.5) | 2.19(1.3-3.4) | 9.54(8.7-10.5) | 18.4(16.4-20.8) | 1.95(1.6-2.4) |
| F | 60-70y | 8072 | 17.1(13.4-21.6) | 2.47(1.5-3.7) | 8.92(8.1-9.8) | 19.7(17.7-22.2) | 2.26(1.9-2.7) |
| F | >70y | 4967 | 16.9(13.5-21.2) | 2.69(1.7-3.8) | 8.39(7.7-9.1) | 21.4(19.2-23.7) | 2.59(2.1-3.1) |
| M | 40-50y | 282 | 13.3(10.5-16.9) | 4.33(2.9-5.9) | 15.3(14.2-17) | 15.3(13.4-17.3) | 1.65(1.2-2) |
| M | 50-60y | 4238 | 13.3(10.4-16.8) | 4.77(3.2-6.5) | 15.1(13.7-16.5) | 15.7(14-17.9) | 1.76(1.4-2.2) |
| M | 60-70y | 7044 | 13.2(10.5-16.5) | 5.08(3.5-6.8) | 14.0(12.8-15.2) | 17.2(15.2-19.4) | 2.13(1.7-2.6) |
| M | >70y | 5976 | 13.0(10.4-16.1) | 5.17(3.6-6.8) | 12.9(11.8-14) | 18.7(16.6-21) | 2.51(2-3.1) |

IMAT, intramuscular adipose tissue. IQR, interquartile range. L, liters. SAT, subcutaneous adipose tissue. SM, skeletal muscle. SMFF, skeletal muscle fat fraction. VAT, visceral adipose tissue.

**Table S5: Baseline characteristics - UKB testing cohort for evaluation of association between body composition and outcomes**

| **Characteristic** | **Overall**, N = 34,638 | **Female**, N = 18,267 | **Male**, N = 16,371 |
|---|---|---|---|
| Age (y) | 64.9 ± 7.8 | 64.3 ± 7.7 | 65.5 ± 7.9 |
| BMI (kg/m$^2$) | | | |
| < 25 | 16,576/34,638 (48%) | 10,015/18,267 (55%) | 6,561/16,371 (40%) |
| 25-29.9 | 13,126/34,638 (38%) | 5,675/18,267 (31%) | 7,451/16,371 (46%) |
| ≥ 30 | 4,936/34,638 (14%) | 2,577/18,267 (14%) | 2,359/16,371 (14%) |
| Race (White) | 33,539/34,549 (97%) | 17,692/18,228 (97%) | 15,847/16,321 (97%) |
| Weekly Alcohol Consumption | 24,644/34,371 (72%) | 11,894/18,110 (66%) | 12,750/16,261 (78%) |
| History of Hypertension | 4,495/34,638 (13%) | 1,946/18,267 (11%) | 2,549/16,371 (16%) |
| Ever smoked | 13,893/34,482 (40%) | 6,766/18,172 (37%) | 7,127/16,310 (44%) |
| History of Cancer | 4,053/34,327 (12%) | 2,243/18,089 (12%) | 1,810/16,238 (11%) |

| **Characteristic** | **Overall**, N = 34,638 | **Female**, N = 18,267 | **Male**, N = 16,371 |
|---|---|---|---|
| Incident Diabetes | 657/34,638 (1.9%) | 229/18,267 (1.3%) | 428/16,371 (2.6%) |
| Follow-up time (y): Diabetes | 4.75 (IQR 3.89-6.08) | 4.76 (IQR 3.91-6.11) | 4.74 (IQR 3.86-6.07) |
| Incident MACE | 575/34,638 (1.7%) | 181/18,267 (1.0%) | 394/16,371 (2.4%) |
| Follow-up time (y): MACE | 4.76 (IQR 3.90-6.12) | 4.77 (IQR 3.92-6.13) | 4.75 (IQR 3.87-6.09) |
| All-cause Death | 573/34,638 (1.7%) | 210/18,267 (1.1%) | 363/16,371 (2.2%) |
| Follow-up time (y): All-cause Death | 4.78 (IQR 3.93-6.15) | 4.79 (IQR 3.93-6.15) | 4.78 (IQR 3.92-6.15) |

**Table S6: Median absolute deviation of body composition GAMs**

| | MAD | | | | |
|---|---|---|---|---|---|
| **Sex** | **SAT (L)** | **VAT (L)** | **SM (L)** | **SMFF (%)** | **IMAT (dL)** |
| Female | 4.15 | 0.90 | 0.76 | 1.89 | 0.33 |
| Male | 3.03 | 1.46 | 1.13 | 1.81 | 0.34 |

GAM, generalized additive model. MAD, median absolute deviation. SAT, subcutaneous adipose tissue. SM, skeletal muscle. SMFF, skeletal muscle fat fraction. VAT, visceral adipose tissue.
